# Quantifying the Financial Value of Clinical Specialty Choices and its Association with USMLE Step 1 Scores

**DOI:** 10.1101/2020.03.14.20036087

**Authors:** Pranav Puri, Natalie Landman, Robert K. Smoldt, Denis A. Cortese

## Abstract

**Importance:** The factors influencing medical student clinical specialty choice have important implications for the future composition of the US physician workforce. The objective of this study was to determine the career net present values of US medical students’ clinical specialty choices and identify any relationships between a specialty’s net present value and competitiveness of admissions as measured by US Medical Licensing Examination (USMLE) Step 1 scores.

**Methods:** Net present values were calculated by using results of the 2019 Doximity Physician Compensation report, a survey of 90,000 physicians. Mean USMLE Step 1 scores for matched US allopathic seniors in the 2018 National Resident Matching Program were used as a measure of clinical specialties’ competitiveness of admissions. We calculated a composite measure of net present value and annual work-hours by dividing each specialty’s net present value by the reported average number of hours worked per year.

**Results:** In our analysis, orthopedic surgery had the highest net present value ($10,308,868), whereas family medicine had the lowest net present value ($5,274,546). Dermatology and plastic surgery had the highest mean USMLE Step 1 scores (249 for both), whereas family medicine had the lowest (220). Clinical specialties’ net present values were positively associated with mean USMLE Step 1 scores (Pearson’s r=0.82; *p*<.001).

**Conclusion and Relevance:** In this study, we describe associations suggesting that medical students choose clinical specialties as rational economic agents and that these decisions are mediated by USMLE Step 1 scores. This underscores the importance of titrating and aligning economic incentives to improve the allocation of medical students into clinical specialties

## Introduction

Globally, public opinion polls rank medicine as the most respected and trusted profession.^1^ Physicians are uniquely positioned within society to alleviate suffering and improve the health of others. Therefore, medicine has long been thought of as a calling, rather than a profession. Psychologists Dik and Duffy describe a calling as a career that has an external summons, provides a sense of meaning or purpose, and is used to help others in some capacity.^2^ In addition, physicians who identify with medicine as a calling are less likely to experience burnout.^3^

On the other hand, a career in medicine is also financially rewarding. Although medical school tuitions and physician debt burdens have increased markedly in recent years, physicians remain among the highest paid professionals.^4^ From an economic perspective, a student’s decision to pursue a medical education can be considered an investment in human capital. Just as a firm’s physical capital comprises buildings and machines, a physician’s human capital comprises medical knowledge and clinical skills. If medical education is an investment in human capital and medical students are rational economic agents, then standard economic theory predicts that medical students will maximize returns on their investment in the same way that firms maximize returns on physical capital.

Prior research has evaluated careers in medicine as investments in human capital. Marcu et al^5^ and Doroghazi and Alpert^6^ have shown that a medical degree confers considerable economic benefit, despite high upfront costs. However, physician earnings vary broadly among clinical specialties. In 2019, the average salary of a neurosurgeon was $617,000, whereas the average salary of a family medicine physician was $242,000.^7^ Similarly, the duration of training also varies widely among clinical specialties. Neurosurgeons complete 7 years of postgraduate medical education, whereas family medicine physicians complete 3 years.^8^ Therefore, the financial investment in a career in neurosurgery is markedly different from that of a career in family medicine.

However, if medical students are primarily motivated by a calling, then they likely will choose clinical specialties on the basis of their academic interests or passions. If medical students select clinical specialties purely on the basis of their interests and those interests are diverse, the investment value of a clinical specialty should not be associated with the competitiveness of admission into the field. For example, if neurology is no less “interesting” than orthopedic surgery, admissions for orthopedic surgery programs should be no more competitive than for neurology programs, even if a career in orthopedic surgery is a more financially rewarding investment.

In the United States, graduating medical students apply to residency programs through the National Resident Matching Program (NRMP). During the past 10 years, the NRMP has become increasingly competitive,^9^ with the average number of residency program applications per US medical graduate increasing from 32 to 60.^10^ In addition, because of the increased adoption of pass/fail grading, US Medical Licensing Examination (USMLE) Step 1 scores are the only non-demographic continuous variable by which residency program directors can screen applicants.^11^ Faced with a growing number of applications, residency program directors have increasingly filtered applicants by Step 1 score, and in a 2018 national survey of program directors, Step 1 score was the most commonly cited factor in choosing candidates to interview.^11^ On February 12^th^ 2020, the National Board of Medical Examiners (NBME) and the Federation of State Medical Boards (FSMB) announced that USMLE Step 1 will transition from a three digit numeric score to a pass/fail outcome, potentially as soon as 2022.^12^ However, until this change takes effect, program directors will likely continue to use USMLE Step 1 scores to filter applicants.

Therefore, the existing literature and trends described above raise the question: do medical students choose clinical specialties as a calling or as an investment in human capital?^13,14^ Further, how is this decision influenced by USMLE Step 1 performance? Although previous studies have attempted to answer these questions by using surveys, stated preferences often differ considerably from the actual choices made by individuals.

In this study, we calculated the investment (net present) values of careers in various clinical specialties. We then evaluated the associations among these net present values (NPVs), annual work-hours, and competitiveness of admissions as measured by USMLE Step 1 scores. Our aim was to identify relationships, which may or may not be causal, to generate hypotheses and facilitate further discussion on these dimensions of medical education.

## Methods

We restricted our analysis to specialties that offered at least 100 positions in the 2018 Accreditation Council for Graduate Medical Education match^9^ and estimated the investment value of careers in various clinical specialties by using a NPV method.^5^ NPVs are frequently used by businesses to project and compare the profitability of different investment options.^15^ The concept of NPV is best explained through an example. Suppose a firm needs to make a payment of $100,000 exactly 2 years from now. If a bank offered the firm a 5% interest rate on deposits, then the firm could deposit $90,702.95 ($100,000/1.05^2^) today, and in exactly 2 years, they would have a balance of $100,000. Thus, the net present value of the payment is $90,702.95.

We calculated NPVs by assuming that students enter medical school at age 24 years (the mean age of matriculating allopathic medical students^16^) and graduate from medical school at age 28 years. Residency training lengths were determined from the Washington University St. Louis residency roadmap application.^8^ We assumed a postgraduate year (PGY) 1 salary of $55,000 with a 3% straight-line increase in annual salary for the duration of the training period. Our assumed PGY 1 salary is comparable to the median PGY 1 salary reported in the AAMC 2018 Debt Fact Card.^16^ We conservatively assumed a 3% annual salary growth rate, as this corresponds to a rate of 1% above inflation. We assumed that upon completion of residency, physicians earned their specialty’s national average salary, as reported by the 2019 Doximity Physician Compensation survey of 90,000 physicians.^7^ From this point through retirement at age 65 years, we assumed a 3% straight-line salary increase for the duration of the physician’s career. We assumed a discount rate of 5% throughout the analysis.

We explore the relationship between NPV and admissions competitiveness across clinical specialties. We used clinical specialties’ mean USMLE Step 1 scores for matched US allopathic seniors as a proxy for admissions competitiveness. We descriptively plot mean USMLE Step 1 scores against NPV and estimate correlation using a Pearson’s correlation.

We then analyze how this relationship is affected by accounting for a clinical specialties’ annual work-hours. We cite clinical specialty annual work-hours reported by Leigh et al,^17^ who surveyed 6,381 physicians to calculate a composite measure of NPV and work-hours. This measure was calculated by dividing each specialty’s NPV by the reported average number of hours worked per year. We illustrate the association between mean USMLE Step 1 scores and annual work-hours weighted NPV and calculate a Pearson’s correlation.

All analyses were carried out using JMP, version 14 and evaluated at a significance level of 0.05. This study was exempt from IRB review since we utilized publicly available datasets that do not contain patient health information

## Results

The results of our analysis are summarized in Table 1. Orthopedic surgery had the highest NPV ($10,308,868), whereas family medicine had the lowest NPV ($5,274,546). Differences in NPV were driven by differences in average annual salary and by duration of training--physicians with longer training periods receive postgraduate salaries for longer periods than their peers in shorter training programs. Dermatology had the highest annual work-hours−weighted NPV (4,388 [$/annual work-hours]), whereas family medicine had the lowest weighted NPV (2,190 [$/annual work-hours]). Dermatology and plastic surgery had the highest mean USMLE Step 1 scores (both 249), whereas family medicine had the lowest mean score (220).

**Table 1.** Summary Statistics

| **Specialty<sup>a</sup>** | **2019**<br>**USMLE**<br>**Step 1 Score,**<br>**mean** | **NPV** | **Annual**<br>**Work-**<br>**Hours** | **Annual Work-**<br>**Hours–Weighted**<br>**NPV, \$/annual**<br>**work-hours** | **2019**<br>**Average**<br>**Salary** | **2019**<br>**Matched**<br>**Graduates**<br>**, No.** |
| --- | --- | --- | --- | --- | --- | --- |
| Dermatology | 249 | \$9,088,220.69 | 2,154 | 4,219.23 | \$450,000 | 497 |
| Plastic surgery | 249 | \$7,724,423.85 | 2,471 | 3,126.03 | \$433,000 | 172 |
| Orthopedic Surgery | 248 | \$10,308,868.00 | 2,715 | 3,797.00 | \$526,000 | 752 |
| Otorhinolaryngology | 248 | \$7,578,424.84 | 2,524 | 3,002.55 | \$398,000 | 328 |
| Radiation Oncology | 247 | \$9,198,312.29 | 2,463 | 3,734.60 | \$486,000 | 179 |
| Neurosurgery | 245 | \$10,192,309.02 | 2,770 | 3,679.53 | \$617,000 | 231 |
| Radiology<sup>b</sup> | 240 | \$8,149,067.01 | ... | ... | \$429,000 | 1,099 |
| General surgery | 236 | \$7,670,463.90 | 2,826 | 2,714.25 | \$403,000 | 1,432 |
| Internal medicine | 233 | \$5,740,034.35 | 2,609 | 2,200.09 | \$264,000 | 8,278 |
| Emergency medicine | 233 | \$7,263,452.26 | 2,205 | 3,294.08 | \$336,000 | 2,458 |
| Anesthesiology<sup>b</sup> | 232 | \$8,199,759.92 | ... | ... | \$405,000 | 1,827 |
| Neurology | 231 | \$6,185,915.53 | 2,415 | 2,561.46 | \$303,000 | 874 |
| Obstetrics and<br>gynecology | 230 | \$6,817,709.85 | 2,778 | 2,454.18 | \$335,000 | 1,392 |
| Pediatrics | 227 | \$4,872,532.48 | 2,212 | 2,202.77 | \$223,000 | 2,867 |
| Psychiatry | 226 | \$5,751,556.94 | 2,209 | 2,603.69 | \$281,000 | 1,720 |
| Physical medicine<br>and rehabilitation | 225 | \$6,422,838.40 | 2,157 | 2,977.67 | \$315,000 | 459 |
| Family medicine | 220 | \$5,274,545.54 | 2,500 | 2,109.82 | \$242,000 | 3,827 |

Abbreviations: NPV, net present value; USMLE, United States Medical Licensing Examination.
<sup>a</sup> Pathology was excluded because it was not reported by Leigh et al,<sup>17</sup> nor was it included in the 2019 Doximity Physician Compensation Report.<sup>7</sup>
<sup>b</sup> Leigh et al<sup>17</sup> did not report annual work-hours for radiology and anesthesiology because of high variability.

We observed a positive association between a clinical specialty’s mean USMLE Step 1 score and the clinical specialty’s NPV (Pearson’s r=0.82; *p*<.001) (Figure 1). Similarly, USMLE Step 1 scores were positively associated with the composite measure of NPV and annual work-hours (Pearson’s r=0.79; *p*<.001) (Figure 2).

**Figure 1.**
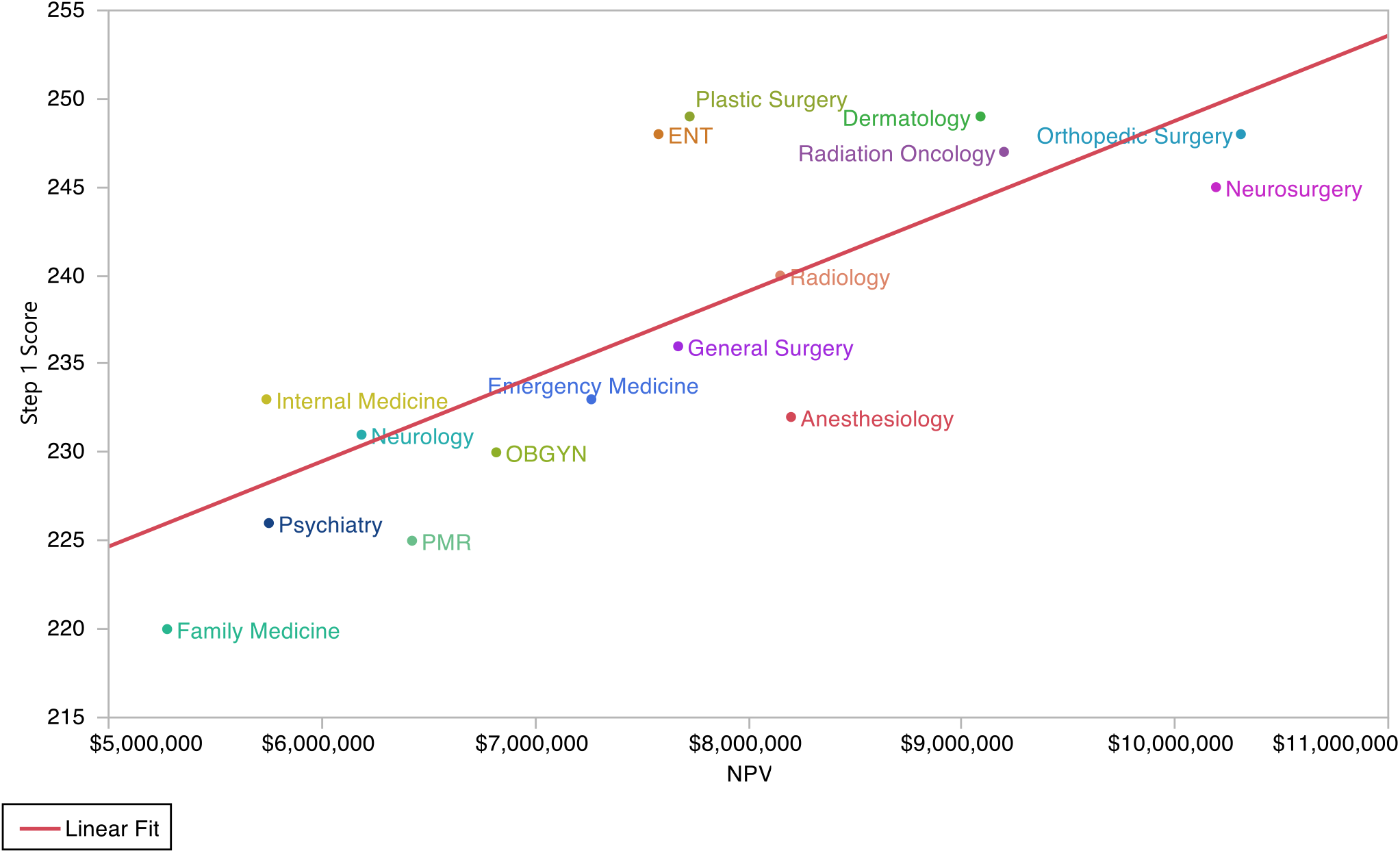
Clinical Specialty Net Present Value (NPV) and Mean United States Medical Licensing Examination (USMLE) Step 1 Score. (Pearson’s r: 0.82, p<0.001) ENT indicates otorhinolaryngology; OBGYN, obstetrics and gynecology; PMR, physical medicine and rehabilitation

**Figure 2.**
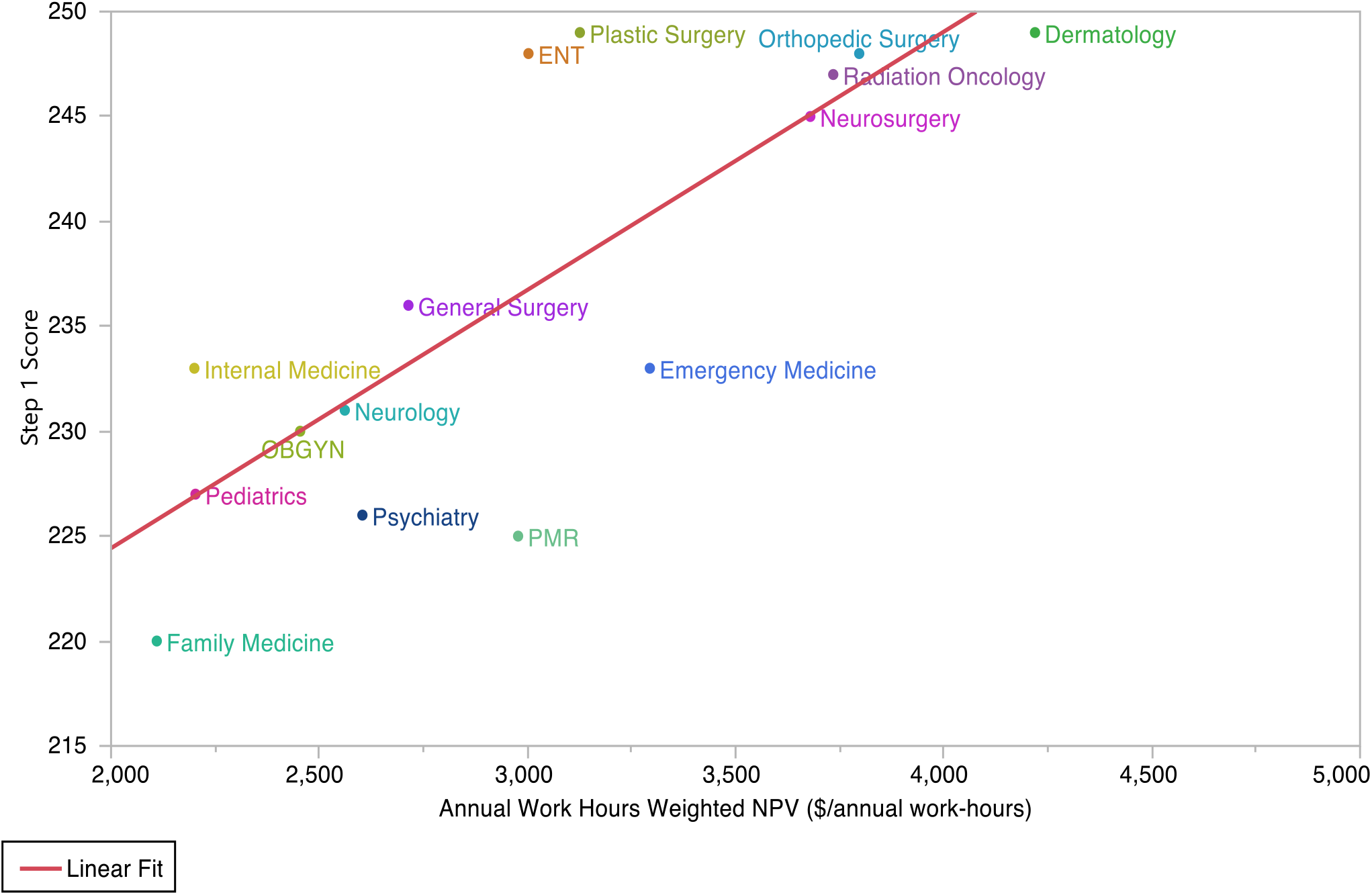
Clinical Specialty Annual Work-Hours−Weighted NPV and Mean USMLE Step 1 Score. (Pearson’s r: 0.79, p<.001) Abbreviations are defined in the Figure 1 legend.

The association between a clinical specialty’s mean USMLE Step 1 score and the specialty’s number of matched graduates was not statistically significant (Pearson’s r= -0.47; *p* = .06). However, after the exclusion of internal medicine as a potential outlier, we observed a statistically significant negative association (Pearson’s r= -0.75; *p* <.001). With the exception of internal medicine, the clinical specialties with the fewest available training positions had the highest mean USMLE Step 1 scores. Similarly, we observed a negative association between clinical specialty NPV and the specialty’s number of matched graduates (Pearson’s r: -0.55, *p*=0.02).

## Discussion

In this study, we calculated the NPV of careers in various clinical specialties and evaluated the associations between NPV, the number of annual work-hours, and USMLE Step 1 scores. Our calculations show wide disparities in the NPVs of clinical specialties, with careers in primary care (family medicine, general internal medicine, pediatrics) having the lowest NPVs. In addition, we identified a positive correlation between a clinical specialty’s NPV and the clinical specialty’s mean USMLE Step 1 score. In other words, the clinical specialties with the highest NPVs were also the most difficult to gain admission into. Similarly, we observed a positive association between a clinical specialty’s annual work-hours−weighted NPV and mean USMLE Step 1 score. These associations, at least in part, suggest that medical students rationally respond to the economic incentives of the residency admissions process.

Medical students spend a substantial portion of time preparing for USMLE Step 1, but this is a rational response to the incentives they face.^19^ To the degree USMLE Step 1 scores serve as gatekeepers to higher NPV specialties, the financial consequences of a student’s USMLE Step 1 score could amount to millions of dollars. Students who obtain high USMLE Step 1 scores are able to clear the cut-off scores applied by program directors and remain competitive for a broad range of clinical specialties. However, students with lower USMLE Step 1 scores may not meet score cut-offs and are screened out from more competitive, high NPV clinical specialties. This restricts students with lower USMLE Step 1 scores to less competitive, lower NPV clinical specialties. Therefore, students with lower USMLE Step 1 scores may not be able to gain admission into the clinical specialties they are genuinely interested in. To the same end, students with higher USMLE Step 1 scores may forego clinical specialties of genuine interest in favor of pursuing higher NPV clinical specialties.

To this end, the NBME and FSMB’s decision to move to pass/fail score reporting for USMLE Step 1 has potential to improve the allocation of physicians into clinical specialties. However, when USMLE Step 1 score reporting transitions to pass/fail, residency program directors will continue to receive increasing numbers of applications, but will be confronted by a paucity of objective, standardized measures to screen applicants. Some have suggested that this will cause USMLE Step 2 Clinical Knowledge scores to assume the current role of USMLE Step 1 scores.^20^ Yet regardless of changes to the admissions process, there will remain wide variations between the NPVs of clinical specialties. This phenomenon plays an important role in medical student specialty choice and creates strong economic incentives for medical students to pursue fields outside of primary care.

This study has several limitations. Because this study used an observational design, we are not able to draw inferences about the causality of the relationships we observe. Confounding factors could influence the relationships we observe. For example, clinical specialties with fewer residency positions had higher USMLE Step 1 scores and higher NPVs. However, the purpose of this study was simply to describe associations between NPV and the competitiveness of residency admissions, which have not been reported in the literature to date. Similarly, we calculated NPVs by assuming physicians earned their clinical specialty’s mean salary, with 3% straight-line growth. However, salaries within clinical specialties can differ broadly, depending on numerous factors such as sub-specialization, geographic location, and practice in an academic or nonacademic setting. In addition, this study did not account for factors that may influence specialty choice, including prestige, outstanding debt, age, race, and gender. Future studies could leverage an experimental design or novel econometric techniques to elucidate the mechanisms underlying the associations described in this paper.

In conclusion, in this study, we describe associations suggesting that medical students choose clinical specialties as rational economic agents and that these decisions are mediated by USMLE Step 1 scores. These associations have important implications for policy-makers, physicians, and medical students alike. The US healthcare system faces a significant and growing shortage of primary care physicians.^21^ At the same time, our calculations show careers in primary care have the lowest NPVs. Therefore, this study underscores the importance of titrating and aligning economic incentives to improve the allocation of medical students into clinical specialties.

## Data Availability

Data is available upon request

https://blog.doximity.com/articles/doximity-2019-physician-compensation-report-d0ca91d1-3cf1-4cbb-b403-a49b9ffa849f

## Acknowledgment

Conflict of interest and financial disclosure: Dr. Landman reports previously investing in Picmonic.

Funding support and role of the sponsor: This study was not externally supported.

Data access and responsibility: Pranav Puri had full access to all the data in the study and take responsibility for the integrity of the data and the accuracy of the data analysis. Pranav Puri takes responsibility for the integrity of the work as a whole, from inception to published article.

## Author contributions

Concept and design: Pranav Puri

Drafting of the manuscript: All authors

Critical revision of the manuscript for important intellectual content: All authors

Statistical Analysis: Pranav Puri

Data Available: Yes

## Abbreviations

NPV: net present value
NRMP: National Resident Matching Program
PGY: postgraduate year
USMLE: US Medical Licensing Examination

